# Effects of Dulaglutide on Ectopic Fat Deposition in Chronic Kidney Disease (CKD): A Pilot and Feasibility Study (GLIMP)

**DOI:** 10.1101/2025.03.19.25324266

**Authors:** Ragibe Gulsah Dilaver, Rengin Elsurer Afsar, Rachelle Crescenzi, Jorge Gamboa, Talat Alp Ikizler

## Abstract

**Introduction:** Patients with chronic kidney disease (CKD) often exhibit ectopic fat accumulation, including intermuscular adipose tissue (IMAT), which is associated with metabolic and muscular dysfunctions. This study aimed to evaluate the effects of dulaglutide, a glucagon-like peptide-1 receptor agonist (GLP-1RA), on reducing IMAT and improving metabolic and physical functions in patients with CKD stage 3-4.

**Methods:** Seven patients were recruited between April 2022 and November 2023. A 12-week dulaglutide (1.5 mg/wk) intervention was conducted with pre-and post-treatment assessments, including magnetic resonance imaging (MRI) for the IMAT evaluation and systemic physical performance battery test (SPPB) for physical performance evaluation. Their body mass indexes (BMI) were calculated and blood samples were analyzed for inflammatory and metabolic markers, including high sensitive C-reactive protein (Hs-CRP), tumor necrosis factor-alpha (TNF-α), interleukin-6 (IL-6), glucose, insulin resistance (IR), total cholesterol, triglyceride, adiponectin, leptin, and leptin-adiponectin ratio (LAR) before and after treatment. Paired t-tests and Mann-Whitney U tests were used for statistical analysis, with significance set at p < 0.05.

**Results:** Out of 58 assessed patients with CKD stage 3-4, 7 were enrolled, with 5 completing the full 12-week dulaglutide treatment. The total 7 people had a mean age of 59 years, mean BMI of 31.4 kg/m², and baseline eGFR of 31.7 mL/min/1.73m². IMAT decreased in 4 patients and increased in 3 patients, with no statistically significant changes overall (p = 0.69). The quadriceps muscle cross-sectional area (CSA) also showed no significant difference (p = 0.73). BMI and serum leptin levels significantly decreased after treatment (p < 0.05), while other inflammatory and metabolic markers, and physical performance scores showed no significant changes. No serious adverse events were reported.

**Conclusions:** This study examined the effects of a 12-week dulaglutide treatment on IMAT accumulation in patients with CKD stage 3-4. While BMI significantly decreased, changes in IMAT were modest and not statistically significant, with potential but unproven clinical and metabolic benefits. Many metabolic and inflammatory markers improved, though not statistically significantly, and physical performance remained unchanged. Muscle CSA and function were maintained, which may alleviate concerns about potential GLP-1RA-induced muscle loss. Dulaglutide was well-tolerated, with minimal side effects. The small sample size and short duration highlight the need for further research.

**Trial Registration:** <u>Name of the Registry</u>: ClinicalTrials.gov

<u>Trial Registration Number</u>: NCT05254418

<u>Date of Registration</u>: 2022-02-01

## INTRODUCTION

Chronic Kidney Disease (CKD) is a significant global health burden worldwide (1). Patients with CKD display many metabolic abnormalities including but not limited to ectopic fat accumulation in multiple organs (2). Ectopic fat accumulation occurs when normal functional adipose tissue reaches its storage capacity, which leads to increased circulating fatty acids and their deposition in non-adipose organs such as liver, muscle and pancreas. This abnormal fat storage can disrupt cellular function and contribute to metabolic diseases, including insulin resistance (IR) and type 2 diabetes (T2D) (3). It is also accompanied by chronic low-grade inflammation, where fat cells in non-adipose tissues release inflammatory markers that further exacerbate the systemic inflammatory state (4).

Intermuscular adipose tissue (IMAT) accumulation, defined as ectopic fat beneath the muscle fascia and between muscle groups, is commonly observed with aging, physical inactivity, and obesity (5). Higher IMAT levels are associated with several metabolic abnormalities, including IR, systemic inflammation, and skeletal muscle mitochondrial dysfunction, as well as functional abnormalities in the skeletal muscle, such as reduced strength, quality, and activation, with decreased gait speed and physical performance (6–8). Muscle function abnormalities and the resulting poor physical performance are common in patients with moderate to advanced CKD, and many studies have shown that IMAT is significantly higher in these patients compared to individuals without kidney disease (9–14).

GLP-1RAs have established benefits for diabetes and obesity and have also demonstrated proven effects in reducing cardiovascular outcomes and albuminuria in diabetic kidney disease patients (15). GLP-1RAs also have demonstrated efficacy in reducing ectopic fat deposition in patients with T2D and obesity (16). These agents have been shown to reduce adipose tissue insulin resistance, leading to lipolysis, reduced inflammatory markers, and increased adiponectin levels (17).

Based on these findings, we decided to perform a small-scale preliminary study before any large-scale research since it is the first study looking at the effects of GLP-1RAs on IMAT accumulation in patient with CKD, other than looking at the kidney or cardiovascular outcomes. We hypothesized that dulaglutide could reduce IMAT accumulation and improve metabolic derangements and physical performance in patients with CKD stage 3-4. We also aimed to test the safety and feasibility of dulaglutide 1.5 mg/wk administration as an adjunct therapy to the standard care of patients with stage 3-4 CKD. To test this hypothesis, we conducted a single-arm, prospective, pilot, and feasibility study of dulaglutide treatment where the subjects served as their own controls.

## METHODS

### Study Population

Participants were recruited from the Vanderbilt University Medical Center (VUMC) Nephrology Clinics between April 2022 and November 2023 (<u>NCT05254418</u>). The primary inclusion criteria for the patients were having stage 3-4 CKD a BMI between 25 – 40 kg/m^2^. Estimated glomerular filtration rate (eGFR) was calculated from creatinine using the Chronic Kidney Disease Epidemiology Collaboration formula. Exclusion criteria included active infectious or inflammatory diseases such as active connective tissue disorders, active cancer, HIV, and liver disease, as well as hospitalization within the last month prior to the study due to a systemic inflammatory process, including conditions like sepsis, severe infection, or acute inflammatory flare-ups. Participants receiving steroids (>5 mg/day) and/or immunosuppressive agents, and individuals using insulin were also excluded. The Institutional Review Boards of VUMC approved the study protocol, and written informed consent was obtained from all study participants. The procedures were conducted under the principles outlined in the Declaration of Helsinki regarding the ethics of human research.

### Study Protocol

Study procedures were conducted at The Vanderbilt Clinics (TVC) and Vanderbilt University Institute of Imaging Science (VUISS). Participants had one screening visit where they underwent a standardized interview, medical records review, and physical examination. The study included two pre-intervention visits and two post-intervention visits, each one month apart, with a 12-week dulaglutide 1.5 mg/wk intervention in between. For each visit, following an 8-hour overnight fasting period, participants presented at VUIIS for Magnetic Resonance Imaging (MRI) and a physical examination, followed by blood draws at TVC. At the second visit, participants began weekly subcutaneous injections of dulaglutide, with the first dose administered during the visit and subsequent doses self-administered over 12 weeks. During the intervention, participants were called biweekly for telephone check-ins and visited monthly to pick up their medication. During biweekly phone calls, participants were asked about any adverse effects. At the end of the twelve weeks, participants returned for the third study visit, and a month later, they had the fourth study visit for follow-up.

### Magnetic Resonance Imaging

Subjects were placed in a 3.0-T MRI scanner (Philips Medical Healthcare, Andover, MA) at each study visit. IMAT was defined as fat located beneath the deep fascia of the muscle and between muscle groups in the mid-thigh section. The IMAT ratio was calculated by dividing the IMAT cross-sectional area by the total muscle cross-sectional area in cross-sectional images of the mid-thigh region between the patella and ischial spine. Images from the thigh were analyzed using a software (MathWorks, Natick, MA) to calculate IMAT. Signal intensities of the pixels in T1-weighted images were used to generate two distinct peaks that differentiate lean tissue from IMAT.

### Physical Function Measurement

The Short Physical Performance Battery test was applied to the participants in each study to evaluate the low extremity functioning in participants (18). The test comprises three main components: the balance test, the gait speed test, and the chair stand test. The balance test involves standing in different positions (side-by-side, semi-tandem, and tandem) for up to 10 seconds each. The gait speed test measures the time it takes to walk a short distance, 3 meters, at a usual pace. The chair stand test assesses lower body strength by timing how long it takes to rise from a chair five times in a row without using the arms. Scores from each component are combined to generate an overall SPPB score ranging from 0 to 12, with higher scores indicating better physical performance.

### Metabolic Markers

After blood was drawn, samples were transported on ice and centrifuged at 3000 rpm for 15 minutes before being stored frozen at −80°C. Plasma fasting glucose concentrations were analyzed using the glucose oxidase method (Glucose analyzer 2; Beckman Coulter, Brea, CA). Biochemistry measurements were analyzed at the VUMC Pathology Laboratory. High-sensitivity C-reactive protein (Hs-CRP) concentrations were measured using the high-sensitivity particle-enhanced turbidimetric UniCel DxI Immunoassay system (Beckman Coulter). Tumor necrosis factor alpha (TNF-α) and interleukin 6 (IL-6) were measured by enzyme-linked immunosorbent assay (R&D Systems, Minneapolis, MN). The Homeostatic Model Assessment for Insulin Resistance (HOMA-IR) was calculated to estimate insulin resistance among the study participants. Total adiponectin was measured using the MILLIPLEX™ MAP Human Serum Adipokine Panel A kit (Millipore, Billerica, MA, USA). Leptin was measured by human radioimmunoassay (RIA) at Vanderbilt’s Hormonal Lab Core. The leptin/adiponectin ratio was determined by dividing the serum leptin level by the serum adiponectin level.

### Statistical Analysis

The characteristics of the participants were presented using the mean and standard deviation for parametric continuous variables, the median and interquartile range for nonparametric continuous variables, and number and percentage for categorical variables. Paired t-tests were used to compare parametric variables, while Mann-Whitney U tests were used to compare non-parametric variables between the groups. The threshold for statistical significance was set at p value <0.05. Analyses were performed using R Studio Version 4.4.2.

## RESULTS

We assessed 58 patients with CKD stage 3-4 for eligibility between April 2022 and November 2023 and enrolled 7 patients. The mean age of the participants was 59 ± 8 years, three were male, all were white, two had T2D, and the mean BMI was 31.4 ± 4.1 kg/m². The baseline eGFR was 31.7 ± 9 mL/min/1.73m². (**Table 1).** Of these, five patients completed the full 12-week treatment period while other two patients, patients GLIMP 3 and GLIMP 6, were able to complete end-of-study visits in the 3rd and 4th weeks, respectively **(Figure 1, 2).** Detailed study schedule can be found in the Supplementary Materials (Table S1).

**Figure 1.**
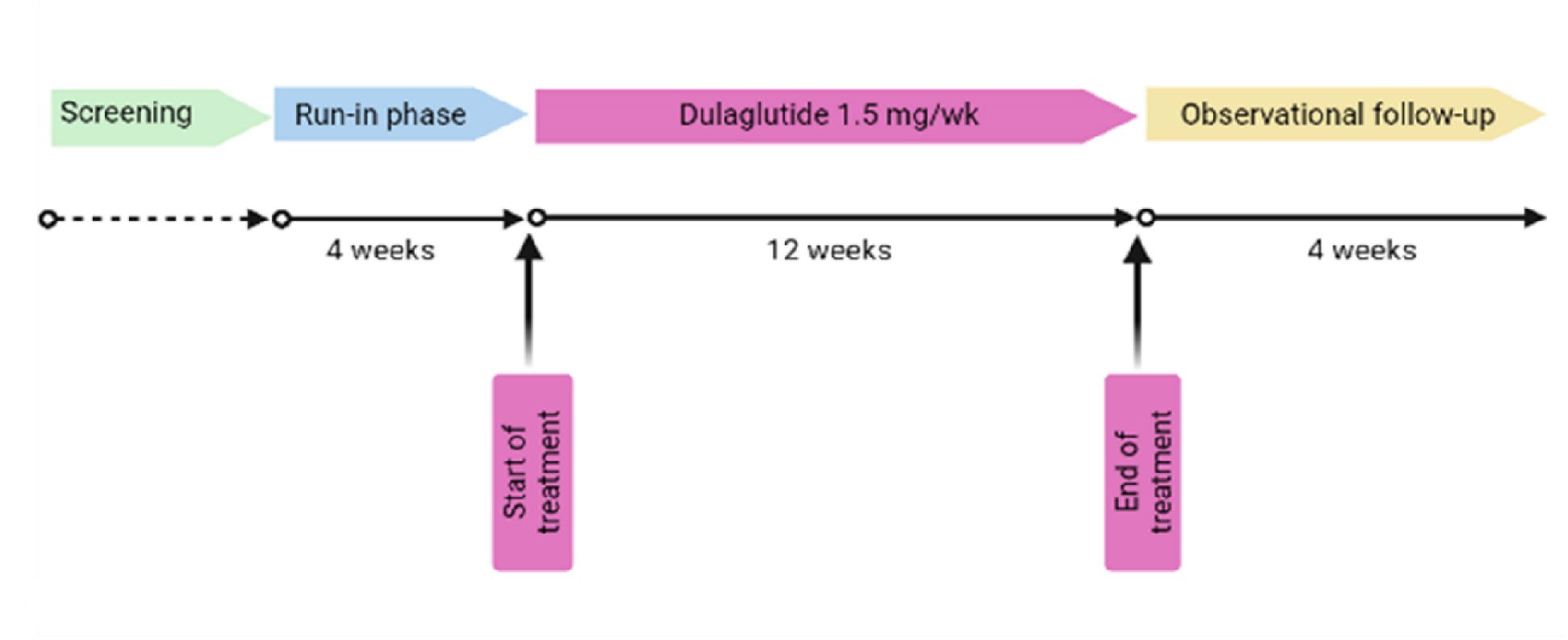
Timeline of the Study Visits and Intervention.

**Figure 2.**
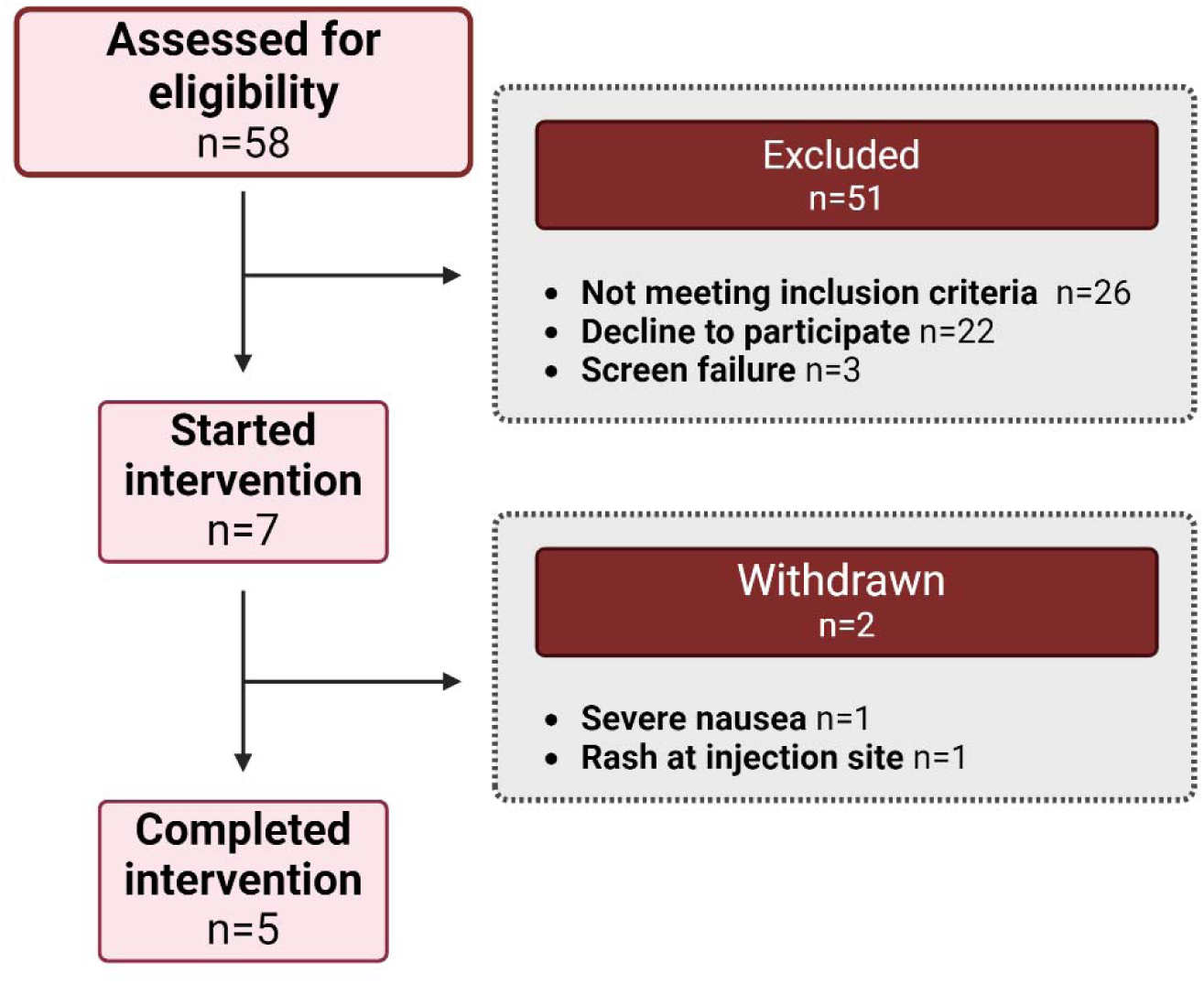
Consort Diagram of the Study Participants.

**Table 1.** Baseline Demographic and Clinical Characteristics of Study Participants.

| **Parameter** | **Total (n = 7)** |
| --- | --- |
| **Age (years)** | 59 ± 8 |
| **Gender, N male (%)** | 3 (42.9%) |
| **Race, N white (%)** | 7 (100%) |
| **eGFR (ml/min per 1.73 m<sup>2</sup>)** | 31.7 ± 9 |
| **BMI (kg/m<sup>2</sup>)** | 31.4 ± 4.1 |
| **Systolic BP (mmHg)** | 114 ± 15 |
| **History of type 2, N (%)** | 2 (28.6%) |

Following the intervention, IMAT decreased in four patients and increased in three patients. The accumulation of IMAT in the thigh muscle group was 0.104 ± 0.041 before the treatment and 0.095 ± 0.033 after treatment (p = 0.69) overall. Among 5 patients who completed the entire treatment, IMAT decreased in 3 patients and increased in 2 patients. The IMAT accumulation was 0.106 ± 0.05 before and 0.099 ± 0.039 after the treatment (p = 0.49), **(Figure 3)**. There was no significant change in the cross-sectional area (CSA) of the quadriceps muscle before and after treatment for all patients in the study (CSA: 223 ± 62 cm^2^ before vs. 220 ± 58 cm^2^ after treatment, p = 0.55), **(Figure 4)**.

**Figure 3.**
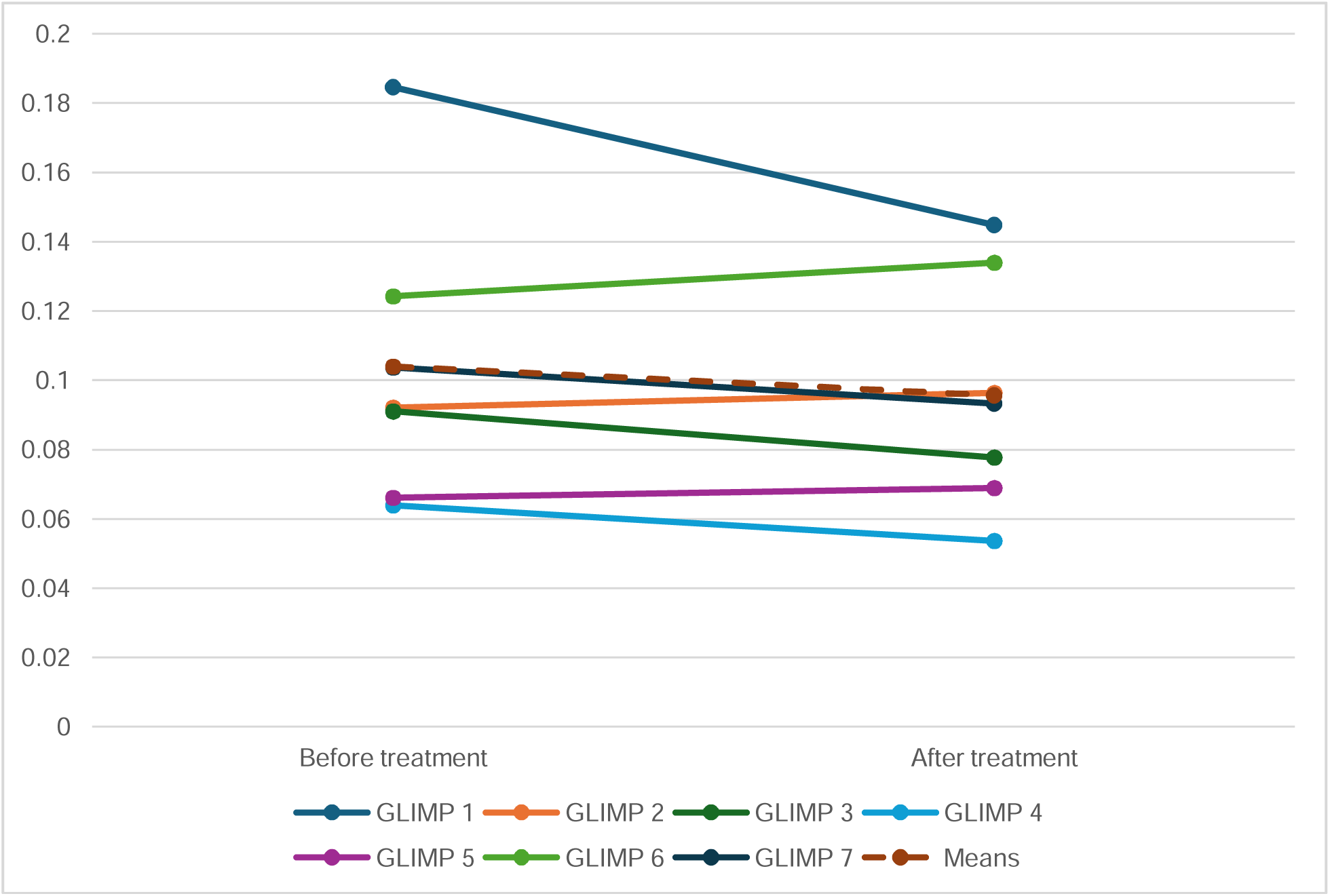
Comparison of IMAT Before and After Treatment.

**Figure 4.**
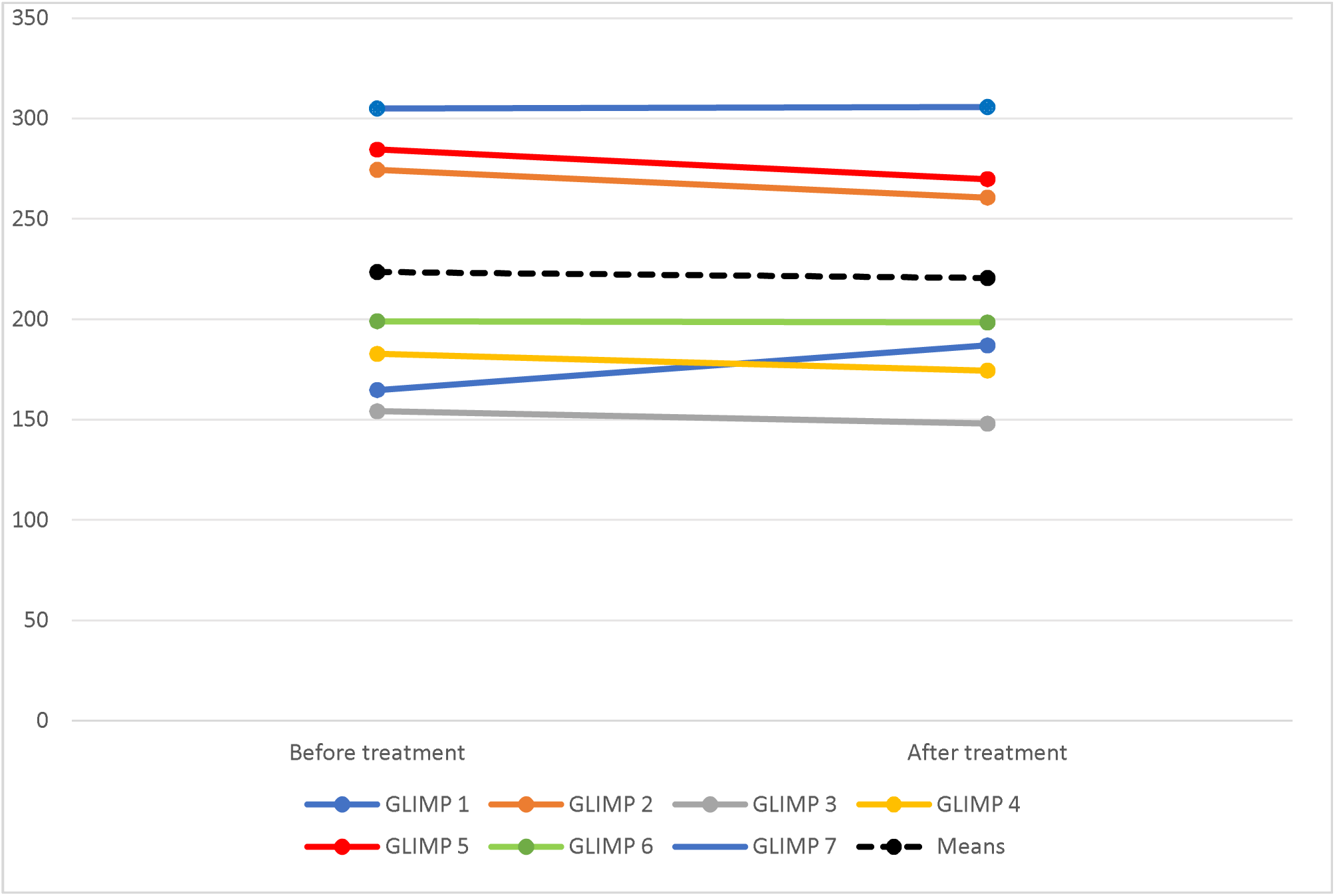
Comparison of Muscle Cross-Sectional Area Before and After Treatment.

**Table 2** provides before and after treatment data for body composition, renal, inflammatory, metabolic markers, and physical performance. Of these variables, body weight, BMI, waist and hip circumferences, blood urea nitrogen, and serum leptin levels decreased statistically significantly after the treatment (p < 0.05). There was no statistically significant difference in any of the other variables, although numerical improvements were consistent among all measured markers. SPPB test scores did not change after treatment (p = 0.88). Detailed complements of SPPB test can be found in Supplementary Materials (table S2).

**Table 2.** Study Parameters Before and After Treatment.

| Parameters | Before treatment | After treatment | p-value |
| --- | --- | --- | --- |
| **Body Composition** |  |  |  |
| **Body Weight (kg)** | 98.94 ± 20.75 | 95.01 ± 18.89 | 0.01 |
| **BMI (kg/m2)** | 35.4 ± 4.1 | 32.3 ± 3.9 | 0.003 |
| **Waist Circumference (in)** | 44.83 ± 5.24 | 43.25 ± 4.96 | 0.03 |
| **Hip Circumference (in)** | 45.58 ± 4.02 | 44.14 ± 3.57 | 0.04 |
| **Waist-to-Hip Ratio** | 0.99 ± 0.11 | 0.97 ± 0.12 | 0.36 |
| **Renal Markers** |  |  |  |
| **eGFR (ml/min/1.73 m<sup>2</sup>)** | 31 ± 7 | 32 ± 10 | 0.93 |
| **Sodium (Na), (mmol/L)** | 139 ± 1.72 | 139 ± 4.21 | 1 |
| **Potassium (K), (mmol/L)** | 4.56 ± 0.54 | 4.64 ± 0.40 | 0.55 |
| **Carbon Dioxide (CO<sup>2</sup>), (mmol/L)** | 22.79 ± 2.31 | 23.86 ± 3.19 | 0.28 |
| **Fasting Plasma Glucose (Glu), (mg/dL)** | 101.64 ± 30.20 | 101.79 ± 30.06 | 0.94 |
| **Blood Urea Nitrogen (BUN), (mg/dL)** | 35.93 ± 12.03 | 29.57 ± 11.52 | 0.02 |
| **Serum Creatinine (Cre), (mg/dL)** | 1.73 (1.62 – 2.3) | 1.82 (1.67 – 2.3) | 0.67 |
| **Calcium (Ca), (mg/dL)** | 9.71 ± 0.41 | 9.90 ± 0.30 | 0.17 |
| **Albumin, (g/dL)** | 4.11 ± 0.31 | 4.19 ± 0.35 | 0.16 |
| **Protein, (g/dL)** | 7.40 ± 0.37 | 7.34 ± 0.42 | 0.7 |
| **Metabolic Markers** |  |  |  |
| **HbA1c (%)** | 6.2 ± 1.2 | 5.6 ± 0.7 | 0.27 |
| **Total cholesterol (mg/dL)** | 183.5 ± 29.8 | 161.4 ± 32.8 | 0.2 |
| **Triglyceride (mg/dL)** | 186.9 ± 61.2 | 168.1 ± 60.6 | 0.31 |
| **Insulin (mIU/L)** | 22.10 (20.70 – 36.81) | 23.16 (17 – 25.58) | 0.71 |
| **HOMA-IR** | 5 (4.25 – 12) | 5 (3.75 – 7.25) | 0.56 |
| **Leptin (ng/mL)** | 21.02 (8.85 – 23.62) | 9.95 (5.08 – 23.62) | 0.04 |
| **Adiponectin (µg/mL)** | 26.97 ± 12.9 | 35.76 ± 18.97 | 0.3 |
| **LAR** | 0.86 ± 0.86 | 0.35 ± 0.25 | 0.15 |
| **Inflammatory Markers** |  |  |  |
| **Hs-RP (mg/L)** | 12.3 ± 6.9 | 10.6 ± 7.1 | 0.66 |
| **TNF-α (pg/mL)** | 5.54 ± 3.39 | 5.33 ± 4 | 0.86 |
| **IL-6 (pg/mL)** | 4.01 (3.185 – 5.87) | 3.37 (1.65 – 6.56) | 0.71 |
| **SPPB test score** | 9 ± 1 | 9 ± 2 | 0.88 |
| **IMAT (fat/muscle ratio)** | 0.104 ± 0.041 | 0.095 ± 0.033 | 0.69 |
| **CSA (cm<sup>2</sup>)** | 223 ± 62 | 220 ± 58 | 0.55 |

Safety data was collected throughout the study period. No serious adverse events were reported. Muscle CSA measured by MRI at the thigh region did not change in the setting of a statistically significant decrease in body weight. One patient experienced nausea and had to discontinue the treatment at the end of week 3. Another patient developed an injection site rash, which led to discontinuation of the treatment at the end of week 4. One patient experienced mild hypoglycemia, which was managed by adjusting their other diabetes medication. No other adverse events were observed.

## DISCUSSION

In this pilot and feasibility study, we investigated the anti-inflammatory and metabolic effects of a 12-week administration of the GLP-1RA dulaglutide and its association with IMAT accumulation in 7 patients with stage 3–4 CKD. We aimed to evaluate whether dulaglutide treatment improves metabolic, functional, and clinical outcomes, as well as its safety and efficacy, in this patient population.

GLP-1RAs have been extensively studied for their ability to reduce major adverse cardiovascular events (MACE) and improve composite renal outcomes in patients with or at risk of atherosclerotic cardiovascular disease (ASCVD), including those with or without diabetes (19). The recently published FLOW trial further confirmed the renal benefits of GLP-1Ras in patients with CKD, highlighting a primary protective effect on kidney function alongside their cardiovascular benefits (20). Notably, the mechanism by which GLP-1Ras exert their kidney beneficial effects is not well-delineated. Further, there are other metabolic effects of GLP-1Ras that extend beyond slowing the progression of kidney disease. Recent studies have demonstrated that patients with heart failure with preserved ejection fraction (HFpEF) experienced significant reductions in heart failure-related symptoms and physical limitations, along with greater weight loss compared to placebo after one year of treatment (22). Moreover, it has been shown that liraglutide reduces the accumulation of visceral adipose tissue, partially independent of weight loss and dosing, which highlights their direct effect on adipose tissue metabolism and systemic benefits beyond kidney function (16,23–25). In this study, we particularly explored the effects of dulaglutide treatment on IMAT and related metabolic parameters in patients with CKD Stage 3. Over the 3-month treatment period, BMI showed a statistically significant reduction, aligning with the expected primary effects of the GLP-1RA class. Although the reduction in IMAT and inflammatory markers, along with increases in adiponectin and decreases in leptin concentrations, were not statistically significant, the observed trends suggest a possible metabolic benefit. Since not every CKD patient benefitted from the perspective of disease progression, these findings suggest that GLP-1RAs should be considered in CKD above and beyond of change sin kidney function.

While ectopic fat infiltration has been linked to adverse metabolic outcomes in various organs, limited data exist regarding its effects on skeletal muscle in CKD, despite the known high prevalence of muscle dysfunction in this population (7). Recent data raise concerns about IMAT’s potential contribution to impaired muscle function and overall physical performance (26). GLP-1RAs have been shown in various studies to enhance physical performance beyond their primary therapeutic effects. For instance, patients with HFpEF treated with semaglutide demonstrated improved 6-Minute Walk Test (6MWT) scores by the end of the treatment period, regardless of diabetes status (22). Similarly, in a study involving T2D patients without cardiovascular disease, liraglutide not only improved myocardial perfusion but also increased the distance covered in the 6MWT (27). This observed improvement in physical performance may be attributed to a complex mechanism that extends beyond weight loss alone involving enhanced muscle insulin sensitivity, mitochondrial function regulation, and reduced local inflammation, which collectively could lead to a reduction in IMAT and improved muscle quality. In our study we did not observe any changes in SPPB test scores, which may indicate that dulaglutide’s impact on physical function in CKD patients is limited, within the short duration of this pilot study. Physical performance in CKD patients can be influenced by multiple factors, including muscle mass, cardiovascular health, and overall physical condition, which might not be significantly altered by short-term metabolic improvements (28). However, from a clinical perspective, patients reported feeling more dynamic and physically active during the treatment period. These subjective reports highlight the need for longer follow-up studies to determine whether the observed improvements translate into objective functional gains over time.

Sarcopenia, defined as decreased muscle mass, function, and strength is very common in patients with advanced CKD (29). Although there are many concerns exist that these medications may exacerbate muscle loss, it is essential to note that concurrent loss of fat and muscle mass may be viewed as a compensatory mechanism due to the elimination of the excess load on the body (30). In our study, we did not see a significant change in muscle cross-sectional area following GLP-1RAs. This finding suggests that the concerns raised in the literature regarding muscle loss associated with GLP-1RAs may not apply to our study population (31). However, we acknowledge the limitations of our study, including the small sample size and short treatment duration. Additionally, all patients were physically active and maintained stable clinical conditions despite their chronic kidney disease, which may have contributed to the preservation of muscle mass. Further research with a larger and more diverse patient population is needed to validate these findings and explore the long-term effects of GLP-1RAs on muscle mass in these populations.

This study is among the first to investigate the effects of pharmacological intervention on IMAT. While the small sample size may have limited the statistical significance of certain findings, the observed trends highlight the potential clinical relevance of IMAT modulation through targeted treatment. The small sample size may have limited power to detect significant differences in key outcomes such as HbA1c, triglycerides, cholesterol, and LAR levels. However, trends like reductions in HbA1c and triglycerides, even without statistical significance, may still hold clinical relevance in the context of metabolic health improvement. Taken together, these changes may suggest an overall metabolic benefit from the intervention despite a short intervention period. Except one patient, our study participants did not mention nausea and vomiting which are common side effects of the medication. This is consistent with the reported side effect profile of GLP-1RAs on the gastrointestinal system. Additionally, one patient experienced an injection site rash. We did not observe dehydration-related adverse effects, which is an important consideration in CKD patients. Overall, the safety profile of dulaglutide in this pilot study was generally acceptable, but the potential for side effects requires careful monitoring, especially in such a vulnerable population.

There are important issues regarding our study that need consideration when interpreting our results. First and foremost, this is a pilot study in a very small cohort with no active comparator. The study duration is relatively short, and we did not have prior data to base on the optimal dosing for the drug of choice. The choice of dulaglutide was based on the availability of incretin mimetics at the time of study initiation, and the results cannot be readily extrapolated to other medications in the class. Even though dulaglutide is a long-acting GLP-1RA that is not cleared by the kidney itself (32), the complex metabolic environment in CKD can affect the efficacy of treatments like GLP-1RAs. Additionally, potential confounding factors, such as variations in baseline metabolic profiles and the presence of comorbid conditions, could have influenced the results. While GLP-1RAs have been effective in reducing ectopic fat and improving metabolic parameters in other groups, the unique metabolic disturbances in CKD may require tailored treatments and longer durations to see significant improvements. Also, newer therapies, such as the dual GLP1-RA/glucose-dependent insulinotropic peptide (GIP) agonist, which are more effective in reducing weight loss, may be more effective in reducing IMAT in patients with CKD (33).

## DECLARATIONS

### 1 – Ethics Approval and Consent to Participate

The Institutional Review Boards of Vanderbilt University Medical Center approved the study protocol, and written informed consent was obtained from all study participants. The procedures were conducted under the principles outlined in the Declaration of Helsinki regarding the ethics of human research.

### 2 – Consent for Publication

All participants provided written informed consent for the publication of their data.

### 3 – Availability of Data and Material

The datasets generated and analyzed during the current study are available from the corresponding author on reasonable request.

### 6 – Authors’ Contributions

RGD contributed to the investigation, formal analysis, and writing of the original draft, as well as the review and editing of the manuscript. REA was responsible for conceptualization, methodology, investigation, and contributed to the review and editing of the manuscript. RC was involved in methodology and the review and editing of the manuscript. JG contributed to conceptualization, methodology, and the review and editing of the manuscript. TAI provided conceptualization, methodology, writing and editing support, as well as supervision throughout the study. All authors read and approved the final manuscript.

## Data Availability

All data produced in the present study are available upon reasonable request to the authors.

## 7 – Acknowledgements

Not applicable.

